# Predicting the distribution of arsenic in groundwater by geospatial machine learning technique in two worst hit districts of Assam, India: a risk to public health

**DOI:** 10.1101/2021.12.30.21268539

**Authors:** Bibhash Nath, Runti Chowdhury, Wenge Ni-Meister, Chandan Mahanta

**Affiliations:** Department of Geography and Environmental Science, Hunter College of the City University of New York, NY 10021, USA; Department of Geological Sciences, Gauhati University, Guwahati, India; Department of Civil Engineering, Indian Institute of Technology Guwahati, Guwahati, Assam, 781039, India

## Abstract

Arsenic (As) is a well-known human carcinogen and a significant chemical contaminant in groundwater. The spatial heterogeneity in the distribution of As in groundwater makes it difficult to predict the location of safe areas for tube well installations for consumption and agricultural use. Geospatial machine learning techniques have been used to predict the location of safe and unsafe areas of groundwater As contaminations. Here we used a similar machine learning approach to determine the risk and extent of As >10 μg/L in groundwater at a finer spatial resolution (250m × 250m) in two worst-hit districts of Assam, India, to advise policymakers for targeted campaigning for mitigation. Random Forest Model was employed in Python environments to predict probabilities of the occurrences of As at concentrations >10 µg/L using several intrinsic and extrinsic predictor variables. The selection of predictor variables was based on their inherent relationship with the occurrence of As in groundwater. The relationships between predictor variables and proportions of As occurrences >10 μg/L follow the well-documented processes leading to As release in groundwater. We identified extensive areas of potential As hotspots based on the probability of ≥0.7 for As >10 µg/L. These identified areas include areas that were not previously surveyed and extended beyond previously known As hotspots. Twenty-five percent of the land area (1,500 km^2^) was identified as a high-risk zone with an estimated population of 155,000 potentially consuming As through drinking water or food cooked with water containing As >10 μg/L. The ternary hazard map (i.e., high, moderate, and low risk for As >10 µg/L) could inform the policymakers to target the regions by establishing newer drinking water treatment plants and supplying safe drinking water.

## 1. Introduction

Groundwater is one of the most critical natural resources which plays a vital role in day-to-day human life and economic development. It is the largest freshwater resource on earth (Foster and Chilton, 2003). However, the pollution of groundwater by naturally occurring arsenic (As) is known to affect fluvial sedimentary aquifers worldwide (Ravenscroft et al., 2009). The As-pollution of groundwater is widespread in the Indian sub-continent, especially in West Bengal (McArthur et al., 2004; Nath et al., 2009) and Bangladesh (Chakraborti et al., 2002; van Geen et al., 2003), where the impact on human health due to the use of contaminated groundwater has remained largely adverse (Smith et al., 2000). The primary exposure route of As to humans is through As-contaminated water, vegetables, and food grains, particularly the consumption of rice (Rahman et al., 2011). Consequently, the World Health Organization (WHO) has set a guideline As a concentration of 10 μg/L in drinking water.

In India and Bangladesh alone, the consumption of groundwater elevated with As has probably killed hundreds of thousands of people prematurely and exposed millions more to a range of ailments (Flanagan et al., 2012). Arsenic in groundwater and its clinical manifestation was first reported in West Bengal, India, in 1984 and later characterized as the “greatest mass poisoning in human history” by WHO (Saha, 1995). Three decades later, a survey of drinking water collected in the homes of many rural households documented that 40 million people in Bangladesh alone were still consuming drinking water with As concentrations >10 µg/L (BBS & UNICEF, 2014). This alarming finding reflects failures towards implementing effective intervention strategies. The installation of private shallow wells has continued unabated but without sufficient knowledge of whether the wells are safe for use.

The occurrence of As in the Brahmaputra flood plain (BFP) groundwater, Assam, India has been discovered much later, threatening the health of millions of consumers (Singh, 2004; Borah et al., 2009; Chetia et al., 2011). The study in Bongaigaon and Darrang districts of BFP demonstrated As enrichment in groundwater ranging from 5 to 606 μg/L with 66% of the analyzed groundwaters containing As concentration above the Indian national drinking water standard and WHO guideline value of 10 μg/L (Enmark and Nordborg, 2007). Mahanta et al. (2015) reported As concentrations >10 ug/L in 29% of the total tested wells based on the state-wide survey of 56,180 tube wells. Verma et al. (2016) and Choudhury et al. (2018) reported that the high As concentration in groundwater is associated with thick clay capping at the top of the aquifer. Choudhury et al. (2018) further suggested that the thick clay layer inhibited flushing of the aquifer, which resulted in high As concentrations in groundwater. The sedimentological history of the BFP is quite similar to the As-contaminated regions of Bangladesh. Therefore, conclusions about the source and mobilization mechanism of As found in Bangladesh can be partly applicable for BFP. The aquifer in this region is much less perturbed from agricultural use of groundwater, which was one of the most critical factors for the occurrence of high As in West Bengal and Bangladesh groundwater (DPHE, 2001). The lack of resources has prevented a comprehensive blanket testing of As in groundwater in the state of Assam, India. Therefore, considering hazards to human health and agricultural use posed by As, mapping the extent of the aquifer contaminated with As is much needed.

Since the inhabitants heavily rely on groundwater for their drinking water and agricultural needs. If a model could be developed through establishing a statistical relationship between As occurrences and environmental predictors (intrinsic and extrinsic) would not only advise policymakers but also inform the villagers of the potential status of their wells, whether safe or unsafe for consumption. The development of a systematic machine learning algorithm would help predict the local-scale distribution of As in groundwater. A similar approach has previously been adopted for the states of Gujarat (Wu et al., 2021) and Uttar Pradesh (Bindal and Singh, 2019), as well as the entire of India (Podgorski et al., 2020a; Mukherjee et al., 2021).

This study focused on risk determination using geostatistical machine learning approaches to identify areas elevated in As concentrations in two worst-hit districts of Assam, India, where previous studies show As concentrations in groundwater increased with distance from the river (Fig. 1). The pattern of this As occurrence is the opposite in comparison to the areas downstream in Bangladesh (Choudhury et al., 2018). Such an observed pattern makes this study unique and challenging in delineating the spatial extent of elevated groundwater As concentrations. In this modeling study, we followed previously studied models but conducted them on a smaller area and at a finer spatial resolution to determine the extent of the problem. Such a finer resolution study would allow the model to characterize the groundwater As occurrences with greater certainty, produce a more accurate model prediction, enable the proper implementation of mitigation measures, and achieve greater benefits.

**Figure 1.**
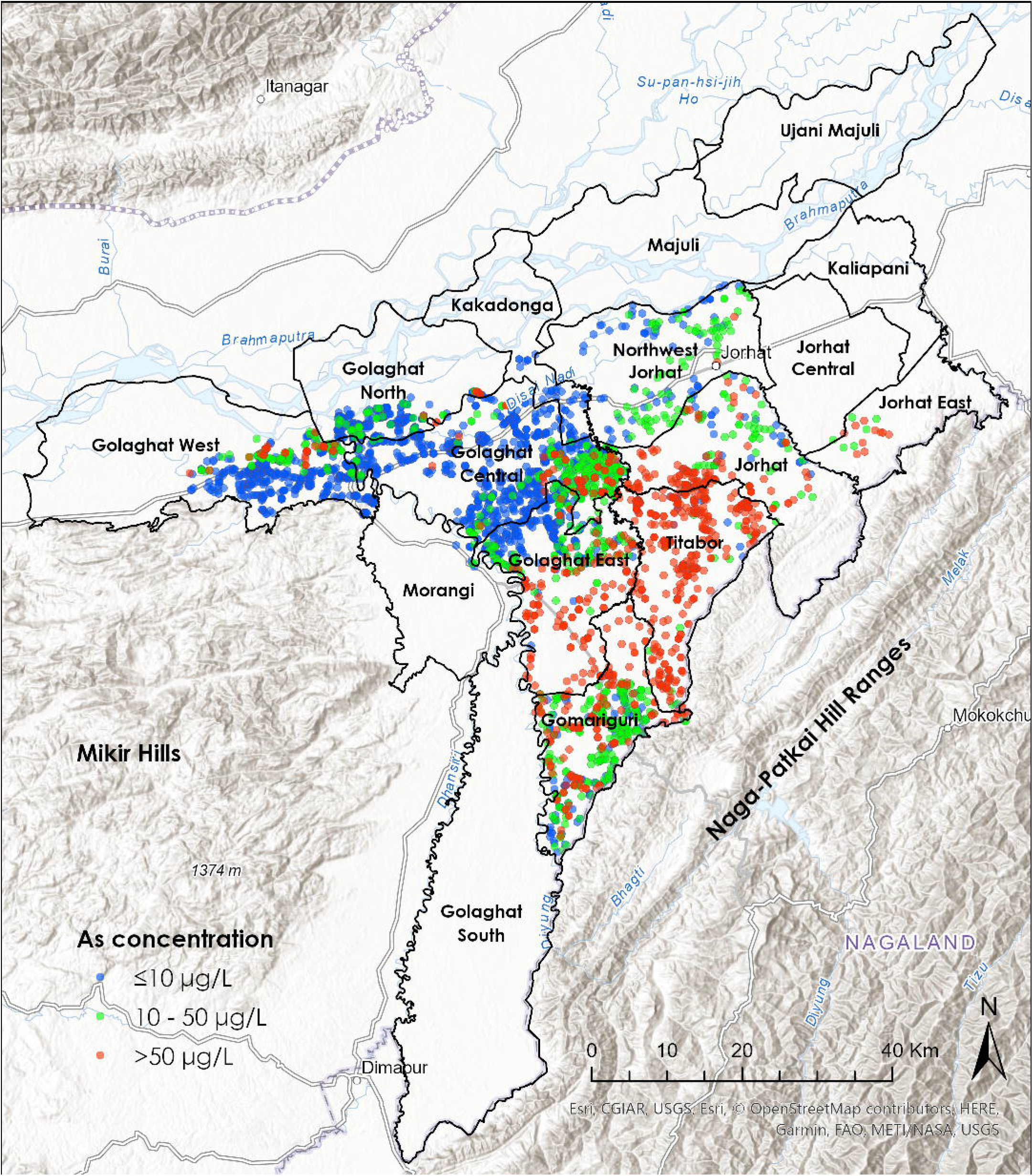
The study area map shows the location of measured As concentrations (n=3,600) in groundwater. The administrative boundaries of sixteen sub-districts (blocks) are also shown.

## 2. Study area for arsenic prediction

The study area is a part of the Brahmaputra River floodplains, which consist of younger and older alluvial sediment depositional environments. The area lies on the southern bank of the Brahmaputra River in Assam, including the river island Majuli. The total area is approximately 6,500 km^2^ (Fig. 1). The area is encircled by the Brahmaputra River to the north, the Naga Patkai Hill range to the south, and the Mikir Hills to the west. The area is marked by high rainfall (average 2,818 mm) and humidity with temperature ranges between 6°C and 38°C. The mean relative humidity is between 92% and 98%. The Brahmaputra River, and its tributaries, Dhansiri, Bhogdoi, and Kakodonga, drain the study area (CGWB, 2008). The alluvial deposits, characterized by light to dark grey colored sands, silts, and clay, are mainly confined to the floodplain areas of the Brahmaputra River and its tributaries (CGWB, 2008).

## 3. Methodology

### 3.1 Random Forest model

The random forest model was implemented in the Python programming language (Pedregosa et al., 2011). The probabilistic relationships between intrinsic and extrinsic predictors and As concentration in groundwater were determined. The random forest model is an ensemble of decision trees. The decision tree algorithm uses a supervised learning method for classification and regression and is non-parametric. The role of a decision tree is to grow by splitting the target variable into consecutive nodes starting from root nodes through decision nodes and ultimately to leaf nodes where the decision is being made about the target. The decision to split the dataset is based on the importance of a predictor variable and its associated conditions. The choice and condition of the predictors to split the datasets are based on how best a predictor decreases the randomness or entropy in the input samples. The best split is selected based on the lowest Gini scores attained after the split.

In a random forest model, each tree uses a different randomly selected subset of predictor variables (typically the square root of the total number of predictor variables) and a random selection with the replacement of data rows (bootstrap aggregating or bagging). Roughly one□third and some data may be repetitive, of the data are used in each tree. Randomness is introduced during the creation of trees to promote uncorrelated forests, avoid multicollinearity among predictors and improve model performance (Ho, 1995). The randomness is created in such a way that each tree is grown using different sample datasets, including a different set of predictor variables. Each tree makes its prediction, and these predictions were then averaged in case of regression or majority votes in case of classification to produce a single result. The random forest model takes care of multicollinearity among predictors because not all variables are used simultaneously in decision trees (Podgorski et al., 2020a).

### 3.2 Target variable: arsenic in groundwater

The measured As concentrations in groundwater were compiled from two sources (Mahanta et al., 2015; Choudhury et al., 2018). These data were generated through a combination of measurements, including in the field using As test kits and in the laboratory using high-resolution inductively coupled plasma mass spectrometry (ICP-MS). For modeling, the data were aggregated to grids of spatial resolution of 250 m by using the geometric mean of concentrations. The extent of the grids was exactly the same as GeoTIFFs of predictor variables so that the grids can be stacked exactly on top of the GeoTIFFs to avoid grids intersecting predictor datasets. The gridded As values were then converted into binary form by assigning all As concentrations meeting the WHO guideline of ≤10 µg/L to zero and all concentrations >10 µg/L to one. This has been done to determine the extent and the total number of people exposed to aid in the decision-making of policymakers or others to identify the priority area for health intervention studies (Podgorski et al., 2020a).

### 3.3 Predictor variables

The occurrence and the spatial extent of groundwater As were modeled through a statistical relationship of the influence of intrinsic and extrinsic predictors and As concentrations within the study area. In total, 26 spatially continuous predictor variables (precipitation, temperature, potential evapotranspiration, aridity, topsoil – and subsoil – organic carbon, sand, silt, clay, cation exchange capacity, pH, bulk density, and coarse fragments, fluvisols, slope, elevation, distance to the river, topographic wetness index and land use/land cover) were used in developing the model. The detailed list of predictor variables and the data sources are given in Table 1. The chosen predictor variables can be broadly classified into climate variables, soil (topsoil and subsoil) characteristics, hydrology, and land use/land cover. The spatial resolution of the predictor variables was mostly 250 m, except for climate variables which are 1,000 m, and distance to the river and topographic wetness index, which are 500 m. We resample those coarser-resolution datasets to finer resolution using spatial interpolation techniques.

**Table 1.** The list of predictor variables was used to develop the random forest model. The data sources and spatial resolution are provided.

| **Predictors** | **Spatial resolution** | **Source** |
| --- | --- | --- |
| Precipitation | 30arc-sec (~1,000m) | Fick and Hijmans, 2017 |
| Temperature | 30arc-sec (~1,000m) | Fick and Hijmans, 2017 |
| Potential evapotranspiration (PET) | 30arc-sec (~1,000m) | Trabucco and Zomer, 2018 |
| Aridity | 30arc-sec (~1,000m) | Trabucco and Zomer, 2018 |
| Topsoil organic carbon content | 250m | Poggio et al., 2021 (SoilGrids) |
| Subsoil organic carbon content | 250m | SoilGrids |
| Topsoil sand content | 250m | SoilGrids |
| Subsoil sand content | 250m | SoilGrids |
| Topsoil silt content | 250m | SoilGrids |
| Subsoil silt content | 250m | SoilGrids |
| Topsoil clay content | 250m | SoilGrids |
| Subsoil clay content | 250m | SoilGrids |
| Topsoil cation exchange capacity | 250m | SoilGrids |
| Subsoil cation exchange capacity | 250m | SoilGrids |
| Topsoil pH | 250m | SoilGrids |
| Subsoil pH | 250m | SoilGrids |
| Topsoil bulk density | 250m | SoilGrids |
| Subsoil bulk density | 250m | SoilGrids |
| Fluvisols | 250m | SoilGrids |
| Topsoil coarse fragments | 250m | SoilGrids |
| Subsoil coarse fragments | 250m | SoilGrids |
| Slope | 30m | Computed from SRTM elevation. |
| Elevation | 30m | Farr and Kobrick, 2000 (SRTM) |
| Distance to river | 15arc-sec (~500m) | Lehner et al., 2008 |
| Land use/land cover | Polygon | Roy et al., 2016 |
| Topographic wetness index | 500m | Hengl, 2018 |

The predictor variables were selected on the basis of their relationships to the process of accumulation and dissolution of As in groundwater (Podgorski et al., 2020a,b; Mukherjee et al., 2021). The soil parameters, estimated at 2.0-m depth, can create geochemical environments favorable for As release. The enriched concentrations of organic carbon in soils favor the development of reducing conditions that result in As release processes. Flat surface topography indicates a low hydraulic gradient, sluggish groundwater flow, and more sediment-water interactions (Nath et al., 2005). Young alluvial sediments, such as high fluvisols probability, indicate conditions similar to the presence of Holocene sediments that have been associated with higher As concentrations (McArthur et al., 2011).

The values of the predictor variables were extracted at the centroid location of each grid pixel containing known (for model development) and unknown (for prediction) As concentrations. During the preparation of the random forest model, the selected environmental predictors were statistically evaluated (Pearson correlation coefficient) and verified the significance (p-value <0.05) of the association to the percentage of grid-averaged As values exceeding 10 µg/L. This was done by organizing the datasets into 12 bins, each containing the same number of observations. The percentage of As measurements in each bin >10 µg/L was then calculated. Sturges’ formula (1 + log2 n) was adopted to determine the optimal number of bins (Sturges, 1926). The statistical significance of the predictors (mean values) in each bin and the percentage of As concentrations >10 µg/L were checked before being included in the model. The dataset was then randomly divided into training (80%) and testing (20%) samples by preserving the ratio of high and low As values through stratified sampling.

### 3.4 Modeling and validation

The random forest model was developed in Python programming language using the “Scikit-learn 1.0.1” package (Pedregosa et al., 2011). The scikit-learn package is efficient and straightforward in predictive data analysis, and the user can tune several hyperparameters to improve the efficiency of the model’s accuracy. We developed the model by growing 1,000 trees using a training dataset. The testing dataset was used to cross-validate the model and determine the accuracy of the model in the prediction of low (≤10 µg/L) and high (>10 µg/L) As concentrations. The probability values of the occurrence of As concentration >10 µg/L in groundwater for the entire study area were determined by applying the model to 18 spatially continuous and statistically significant predictor variables.

Mean decrease in accuracy and mean decrease in Gini scores were computed during model development of each tree grown to evaluate the importance of predictor variables. The mean decrease accuracy and mean decrease Gini score are higher for the more important predictor variables used in the model. Gini node impurity, a measure of misclassification, was calculated for every split based on the probability of samples belonging to a single class. In other words, how much information was gained, randomness dropped, and purity achieved in the samples after each splits. Predictors and the corresponding criterion that provide the best splits in the samples are selected first which is based on the values of the lowest Gini score, i.e., the largest difference in Gini scores before and after the splits. Gini purity indicates the homogeneity of the samples obtained after a split by a specific variable condition (Breiman et al., 1984). The importance of the variable increases with the decrease in Gini impurity. The decrease in accuracy is calculated on out-of-bag (OOB) samples by randomly mixing the data for a particular predictor in the OOB samples. A variable of highest importance if the model suffers accuracy if removing that predictor variable.

The accuracy, sensitivity (i.e., true positive rate), and specificity (i.e., true negative rate) values were also calculated. The cutoff value was used to distinguish between high and low As hazard areas. The value was selected where the sensitivity and specificity of the model become equal. The predictive power of the model was evaluated by using the area under the Receiver Operating Characteristic (ROC) curve. The standard deviations of classification outcome from each tree during the development of the forest were estimated for analyzing the model uncertainty. Low standard deviation indicates greater model certainty in the predicted results. Pearson residuals were also calculated to test model under- and over-predictions.

### 3.5 Estimation of the total area and exposed population

Population data were collected from the WorldPop website for 2020 at a spatial resolution of 1-km (www.worldpop.org). The total land area and the population exposed to elevated As concentrations (>10 µg/L) in drinking water were computed by generating ternary risk maps from the random forest model. Three risk levels were considered based on the cutoff values.

The sensitivity and specificity values were plotted for 100 probabilities between 0 and 1 (Fig. 2). The cutoff point was chosen where sensitivity and specificity values intersect to avoid biases towards either true positive rate or false positive rate (i.e., predicting high or low As concentrations, respectively). Instead of providing two risk levels (low and high hazard areas), we chose three risk levels (high – probability ≥0.7, moderate – probability >cutoff point but <0.7, and low – probability <cutoff point). Such an approach was adopted to identify the priority location for immediate interventions for mitigation and generating community awareness. The hazard area map was then used to calculate the number of at-risk populations in different sub-districts (community development blocks). The total number of people living in the hazard areas in different sub-districts were then multiplied by the modeled probabilities, the proportion of people living in urban and rural areas in each sub-district, and use of untreated groundwater in the study area (0.25, based on the survey results of groundwater filtered through a home-made sand filter).

**Figure 2.**
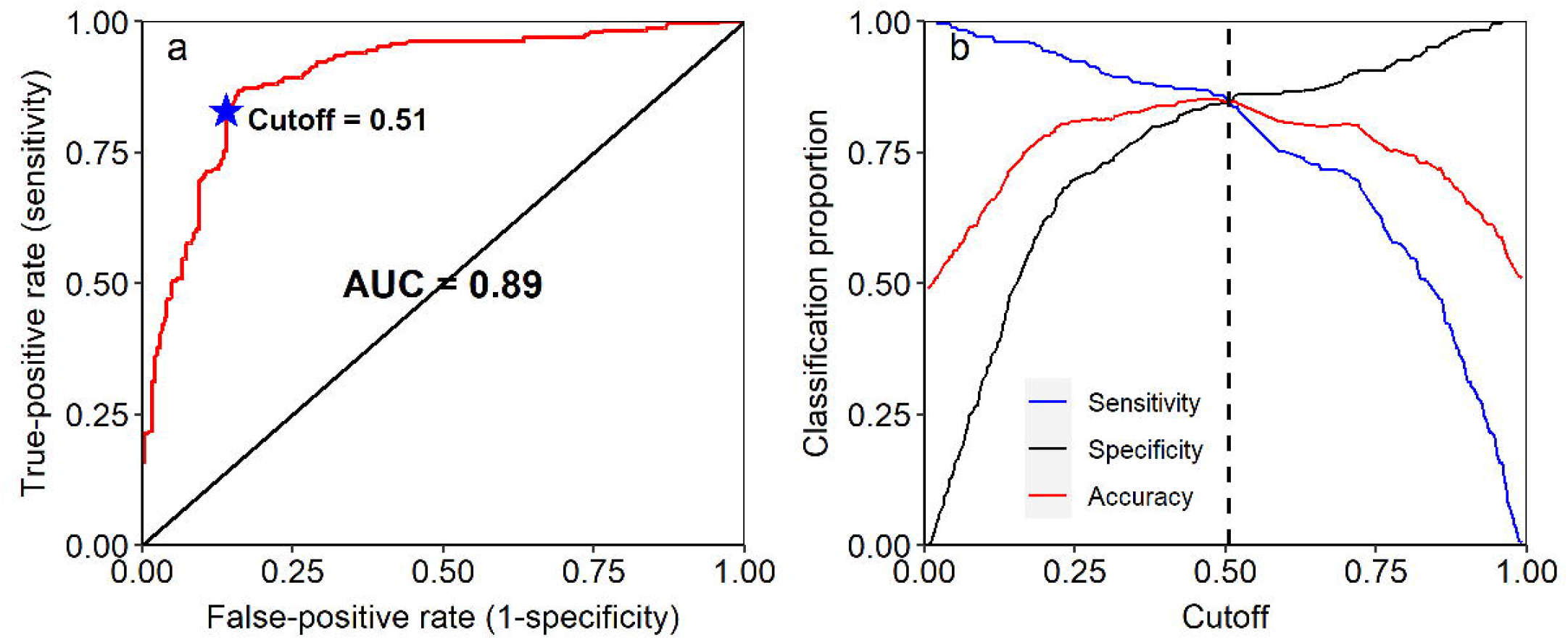
The classification strength. (a) ROC curve with an AUC of 0.89, which indicates the discriminative power of the random forest model. (b) True-positive rate (sensitivity), true-negative rate (specificity), and accuracy against the different cutoff values to identify the best conditions in predicting low and high As concentrations in the study area.

In addition to that, the coverage of piped water supply schemes (PWSS) (Integrated Management Information System, 2021) was also studied to get a better insight into the percentage of people who have access to safe and unsafe water in each sub-districts.

## 4. Results and discussions

### 4.1 Arsenic prediction model outcome

The cross-validation results on the test dataset using the trained random forest model are provided in Table 2. The area under the ROC curve (AUC) is 0.89, which is generally ranges between 0.5 and 1. The values close to 1 indicate a perfect model, while values of 0.5 indicate no better than a random chance occurrence. The AUC value determines the strengths of binary (high and low) classification model prediction. The AUC has been computed by using different probability cutoff values (Fawcett, 2006). The AUC values of 0.89 indicate excellent model performance and are comparable to several other groundwater quality model predictions at both regional scale and country-wide. The AUC value of 0.84 was reported for fluoride prediction in entire India (Podgorski et al., 2018), 0.71–0.83 for As in Gujarat (Wu et al., 2021), and 0.755 for As in Uttar Pradesh (Bindal and Singh, 2019). The test dataset was also used to calculate the model’s overall accuracy (0.85) following 10-fold cross-validation. In addition to that, the accuracy value (0.85) is very close to model accuracy with OOB samples of 0.83. The no information rate is 0.5042 (p-value < 2.0 × 10^−16^), which is significantly low compared to the accuracy obtained in this model. The no information rate is equivalent to an accuracy that could be achieved without a model (Podgorski et al., 2020a). The no information rate is generally proportional to the dominant class of the dataset, i.e., the percentage of As ≤10 µg/L (50.5%) for this study. Cohen’s kappa statistic value is 0.695, indicating stronger reliability and substantial agreement of the model.

**Table 2.** The confusion matrix and statistics of the random forest model were developed based on eighteen significant predictor variables. The probability cutoff value is 0.51.

| **Predicted class** | **Actual class** |  |
| --- | --- | --- |
|  | 0 | 1 |
| 0 | 208 | 36 |
| 1 | 37 | 198 |
| Accuracy | 0.848 |  |
| Cohen's kappa | 0.695 |  |
| Sensitivity | 0.846 |  |
| Specificity | 0.849 |  |
| Positive predicted value | 0.843 |  |
| Negative predicted value | 0.852 |  |
| Prevalence | 0.511 |  |
| AUC | 0.89 |  |

The spatial averaging of the As values from 3,600 locations to 250-m pixels produces high-density data points in the study regions. The grid-averaged 2,400 locations were found to distribute evenly throughout the study area, suggesting any unlikely bias in the model prediction by providing excess weight to the environmental conditions in some areas. Such a situation can occur if the study area is very large with an imbalance in sampling density (Podgorski et al., 2020a). In comparison to other studies, our study site is rather very small and limited to one river basin (Bindal and Singh, 2019; Podgorski et al., 2020a). Therefore, one single random forest model can effectively account for heterogeneity in the geochemical environments and produce a reliable outcome for this study area. This has been made possible using large sets of predictor variables representing climate, land use/land cover, and soil properties used to define the different geochemical environments.

The predicted probability identifies known As-rich areas near the foothills regions of the Naga-Patkai range (Fig. 3a). The model also identifies newer areas and previously unknown areas such as parts of sub-districts Golaghat South, Morangi, Jorhat East, Jorhat Central, and Kaliapani. These areas lack sampling for As measurements, and no public records are available for use in the model. Thus far, the model highlights the advantages of a prediction model over spatial interpolation (such as kriging and IDW) and the usefulness of machine learning techniques that could handle large datasets with many predictors. The predictions can be made in areas without prior knowledge of As testing data. The developed model can be effectively used to prioritize testing in areas where the data is lacking and to confirm potential threats. This study can further benefit from new data from the areas that lack sampling to strengthen the predictive power and accuracy of the model.

**Figure 3.**
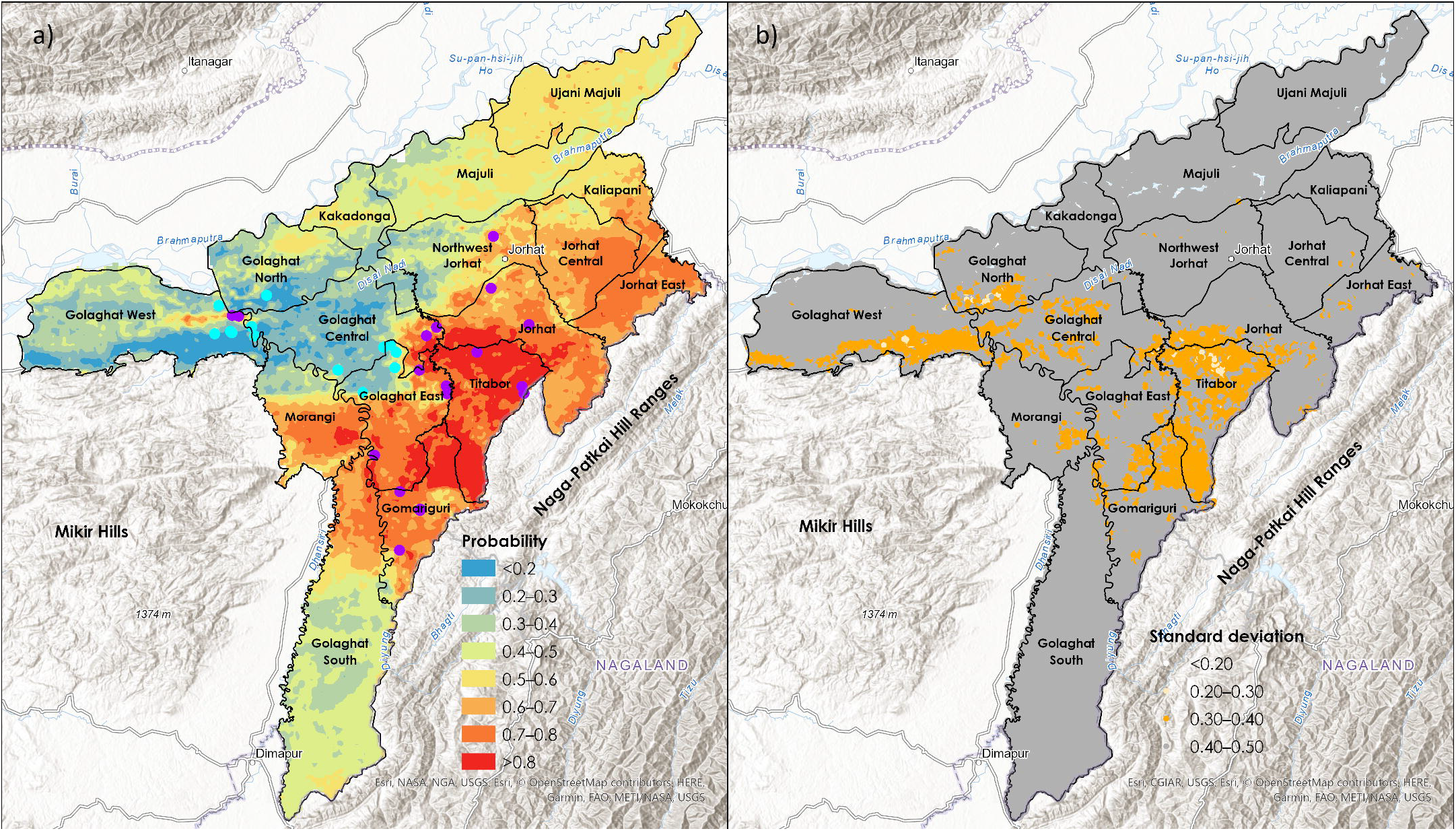
Maps showing the (a) Probability of As >10 μg/L in groundwater in the study area predicted by random forest model. The cyan dots represent grid locations with model under-prediction, and magenta dots represent grid locations of model over-prediction based on Pearson residuals of predicted probability of As >10 μg/L, (b) Standard deviation of predicted class (either 0 or 1) in each grid location.

The standard deviation of the predicted classes for each of the 1,000 decision trees is shown in Fig. 3b. The standard deviation values can be used to assess model uncertainty in the final predicted class. The data showed that the areas with a high predicted probability of As >10 μg/L have a low standard deviation in the predicted class. At the same time, the areas with slightly higher standard deviations are associated with medium to high predicted probability. These areas are in Majuli, Ujani Majuli, and Golaghat South sub-districts, where the sampling is missing. Pearson residuals of the predicted probability of As >10 μg/L were also used to identify the locations of model under- and over-predictions. The results showed that the predictions made by the model show good agreement since the Pearson residuals in most of the grid locations are within the considered range (i.e., Pearson residuals are between <2 and >-2). But only in a handful of locations, the Pearson residuals were either >2 or <-2, suggesting slight over- and under-predictions by the model, respectively (Ayotte et al., 2006). This indicates that the model did a better job in predicting the probability of low and high As concentrations.

### 4.2 Association between environmental predictors and spatial pattern of arsenic

The mean decrease in Gini impurity and mean decrease in accuracy were used to determine the importance of the predictor variables in the random forest model (Fig. 4). The results showed greater importance of subsoil silt and all climate variables (i.e., precipitation, potential evapotranspiration, aridity, and temperature). In addition to that, topsoil silt, elevation, and subsoil clay show strong importance in the model prediction. The least important variables are subsoil and topsoil pH and slope, suggesting least useful in the model.

**Figure 4.**
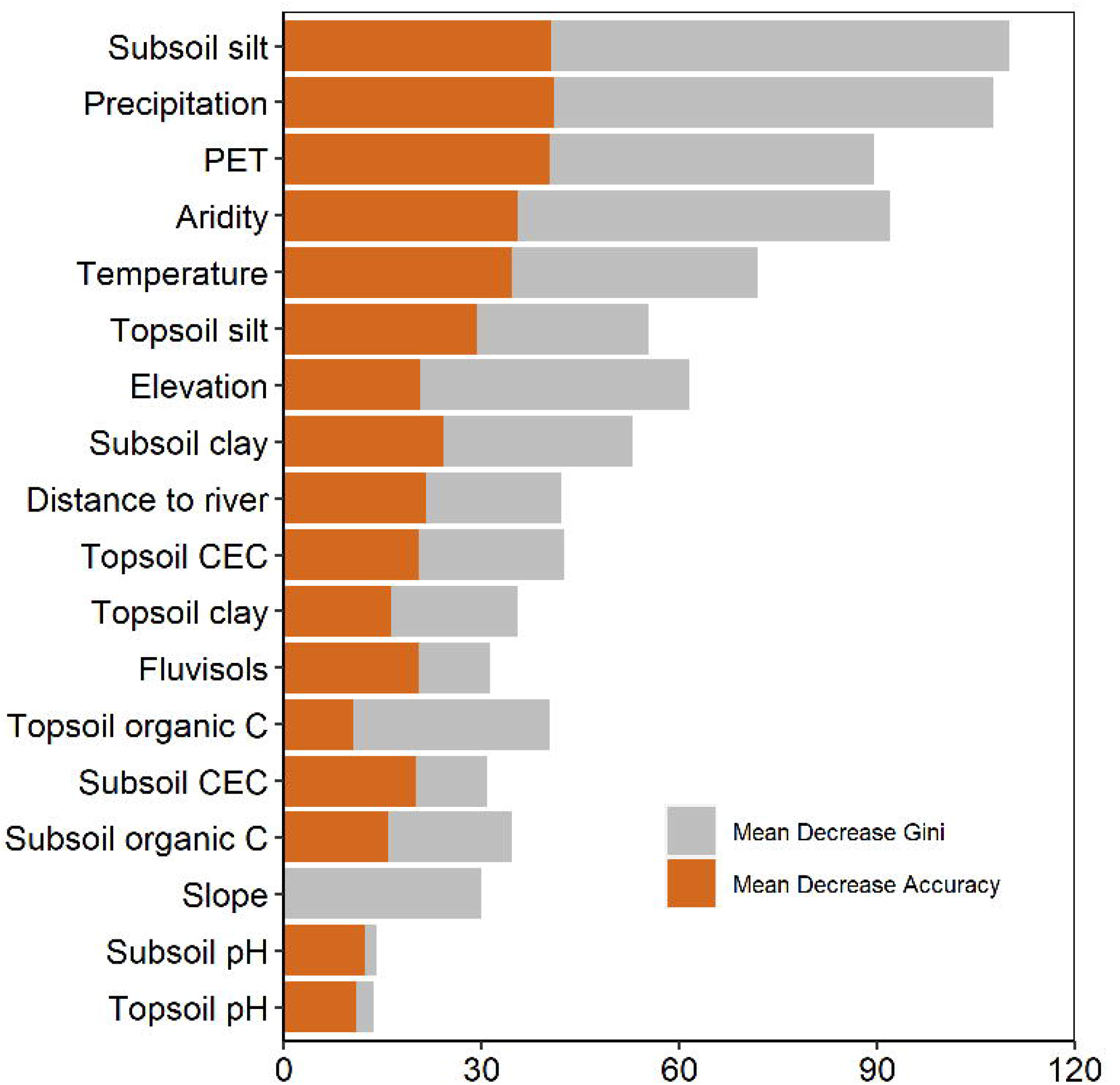
The importance of the predictor variables in relation to mean decrease in Gini and mean decrease in accuracy as calculated by random forest model.

The strength of the relationships between predictor variables (average in each bin) and As concentrations (the percentage of As measurements >10 µg/L) was tested using Pearson correlation coefficient (R) and p-value (95% confidence level) (Fig. 5). The strongest R values were found with topsoil clay, subsoil clay, topsoil silt, subsoil silt, topsoil organic carbon, subsoil organic carbon, topsoil cation exchange capacity, subsoil cation exchange capacity, elevation, and fluvisols (Fig. 5). These variables have R values >0.88 (either positive or negative) and show statistical significance at p-values <0.05. In addition, precipitation, temperature, potential evapotranspiration, and distance to the river also showed stronger relationships with the percentage of As measurements >10 µg/L. Subsoil and topsoil sand have shown well-defined troughs with the lowest proportion of As measurements >10 µg/L coincide mean values of 38 and 34 weight %, respectively (Figs. 5i,j). These variables were also tested in developing the final model since the random forest algorithm is highly effective in capturing nonlinear relationships between predictor and response variables (Ryo and Rillig, 2017).

**Figure 5.**
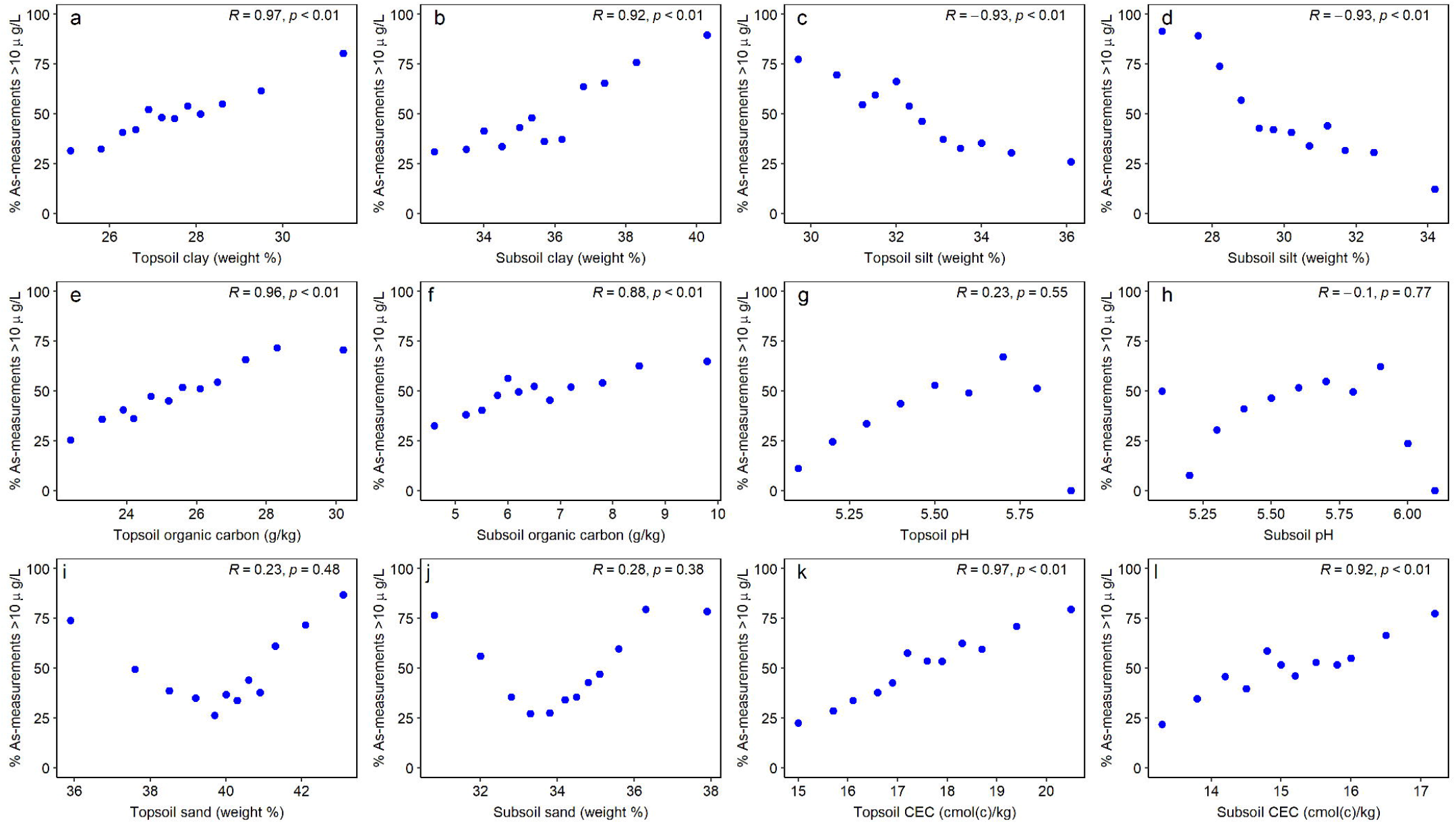
The predictor variables (**a–x**) with percentages of grid-averaged As data exceeding 10 µg/L in 12 equally sized bins and their correlations. Pearson correlations (R) with a statistically significant p-value (95% confidence level) are shown.

Our model is strongly dependent on the subsoil silt content, and a negative relationship with elevated As concentrations signifies the occurrence of high As concentrations in older floodplains. High subsoil silt content was mostly found to localize in the areas close to the current river channel. Higher silt content in soils is indicative of the deposition of fresh sediments by the river (Ahmed et al., 2004). Silt is highly reactive, produced by mechanical weathering; therefore, the presence of high silt content suggests more active sorption sites for As adsorption (Amini et al., 2008). On the contrary, we observed a higher occurrence of subsoil clay further away from the current river channel and found it to be associated with elevated As concentrations. The observation of greater clay fractions and clay capping to greater aquifer depths support leaching of As to groundwater and lack of monsoonal dilution by rainwater. Such an observation increases extensive sediment-water interactions and minimal flushing (Choudhury et al., 2018). Therefore, the composition of the aquifer materials is vital to the occurrences of As in groundwater (Bindal and Singh, 2019).

Our model also shows the high importance of climate variables (Podgorski and Berg, 2020b). Low precipitation and evapotranspiration are associated with a high percentage of As >10 µg/L in tube wells suggesting slower recharge and greater sediment-water interactions. Low precipitation favors reduced flow of groundwater in the aquifer and lack of dilution effects and flushing by infiltrating rainwater and thereby increases the likelihood of As accumulation and release in groundwater over a longer time (Rodríguez et al., 2004). The combined effect of precipitation and evapotranspiration could favor As release due to saturated conditions and forming reducing environments for As release (Podgorski and Berg, 2020b).

The elevation shows moderate importance in our model. However, the relationship between elevation and the percentage of As >10 µg/L is highly significant and is positively correlated. This suggests the occurrences of As away from the river channel instead of the observation in Bangladesh, where low elevation is associated with tube wells containing As concentrations >10 µg/L (Shamsudduha et al., 2008). Interestingly, our previous studies observed the occurrence of brown (oxidized) sands near the river channel, suggesting flushing of the aquifer near the river (Choudhury et al., 2018).

### 4.3 Arsenic-hazard probabilistic zones

The estimated As-risk areas and population exposed were computed based on the probability cutoffs of 0.51 (Fig. 6). The probability of As >10 µg/L was grouped into three categories (high, moderate, and low-risk zones). The high-risk zones are predominantly located south of the Brahmaputra River and beside the Naga-Patkai Foothill ranges. The moderate risk areas are typically extending from high-risk zones toward the river, including the island Majuli. The low-risk zone is mainly located near the river channel, including isolated pockets of moderate-risk zones. In addition to that, a large part of the Golaghat South sub-district is within the low-risk zone, which is bordered by Naga-Patkai hill ranges on the east and Mikir hills on the west. These areas are located at a higher elevation with greater slopes, and thus favorable for greater aquifer flushing and unfavorable for As release to groundwater. The population map shows a cluster of densely populated localities within the high-risk zone (Fig. 6b). Thus, potentially causing a public health concern.

**Figure 6.**
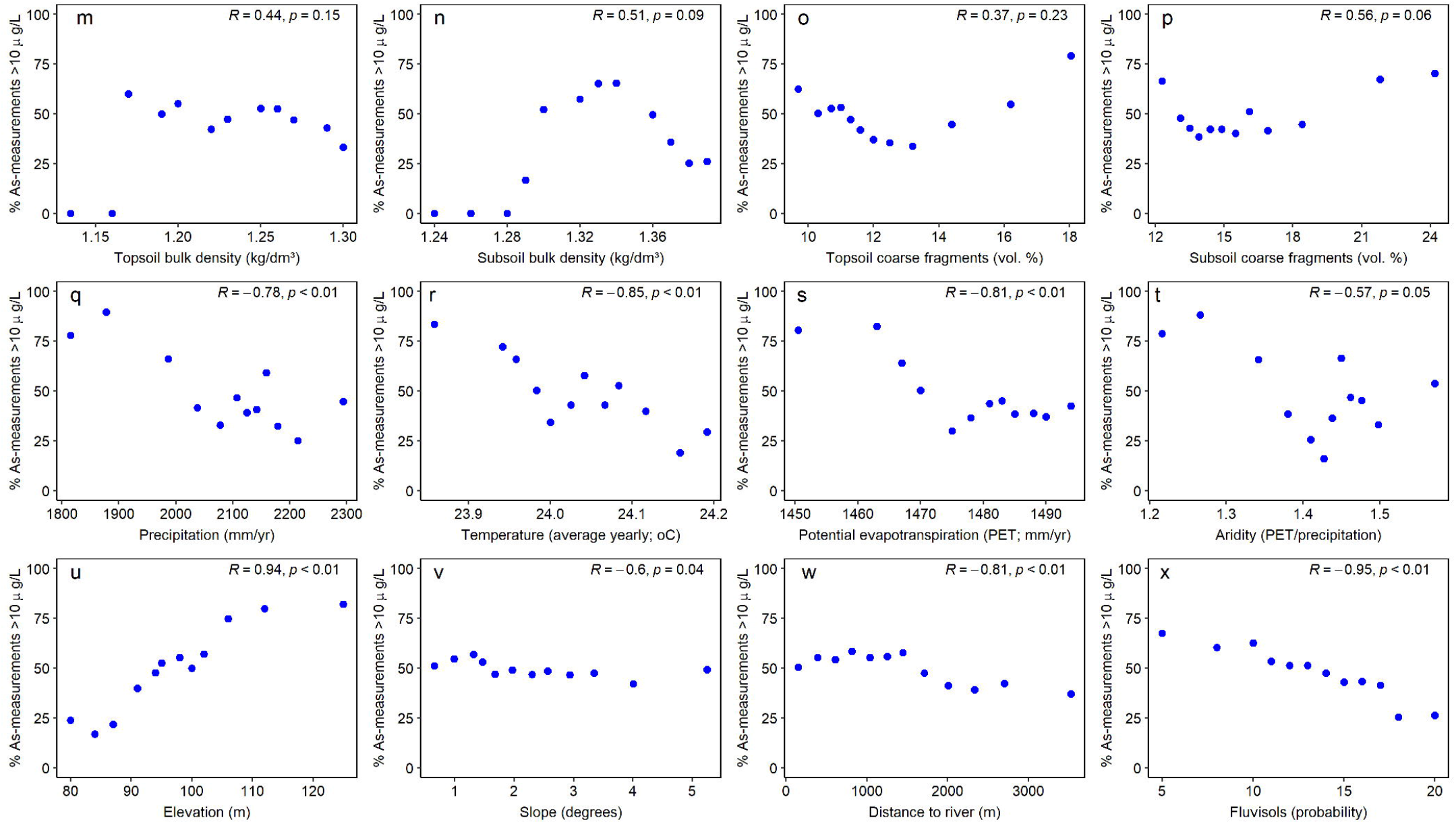
(a) Ternary As hazard (high, moderate, and low) maps based on predicted probability (high risk: >0.7, moderate risk: 0.51-0.7, and low risk: <0.51). (b) The density of population (per Km^2^) living in high-risk areas.

The total populations potentially exposed to As concentrations >10 µg/L are shown for each sub-district (Fig. 7). Based on the probability scores (cutoff scores of 0.51), a total of about 115,000 people in moderate risk areas (probability scores between 0.51 and 0.70) and a further 155,000 people in high-risk areas (probability scores >0.70) may be directly or indirectly exposed to As concentrations >10 µg/L. Most of the exposed population in high-risk areas are living in four sub-districts, i.e., Titabor (a total of 42,000 people), Jorhat (a total of 26,750 people), Golaghat East (a total of 25,500 people), and Gomariguri (a total of 24,000 people).

**Figure 7.**
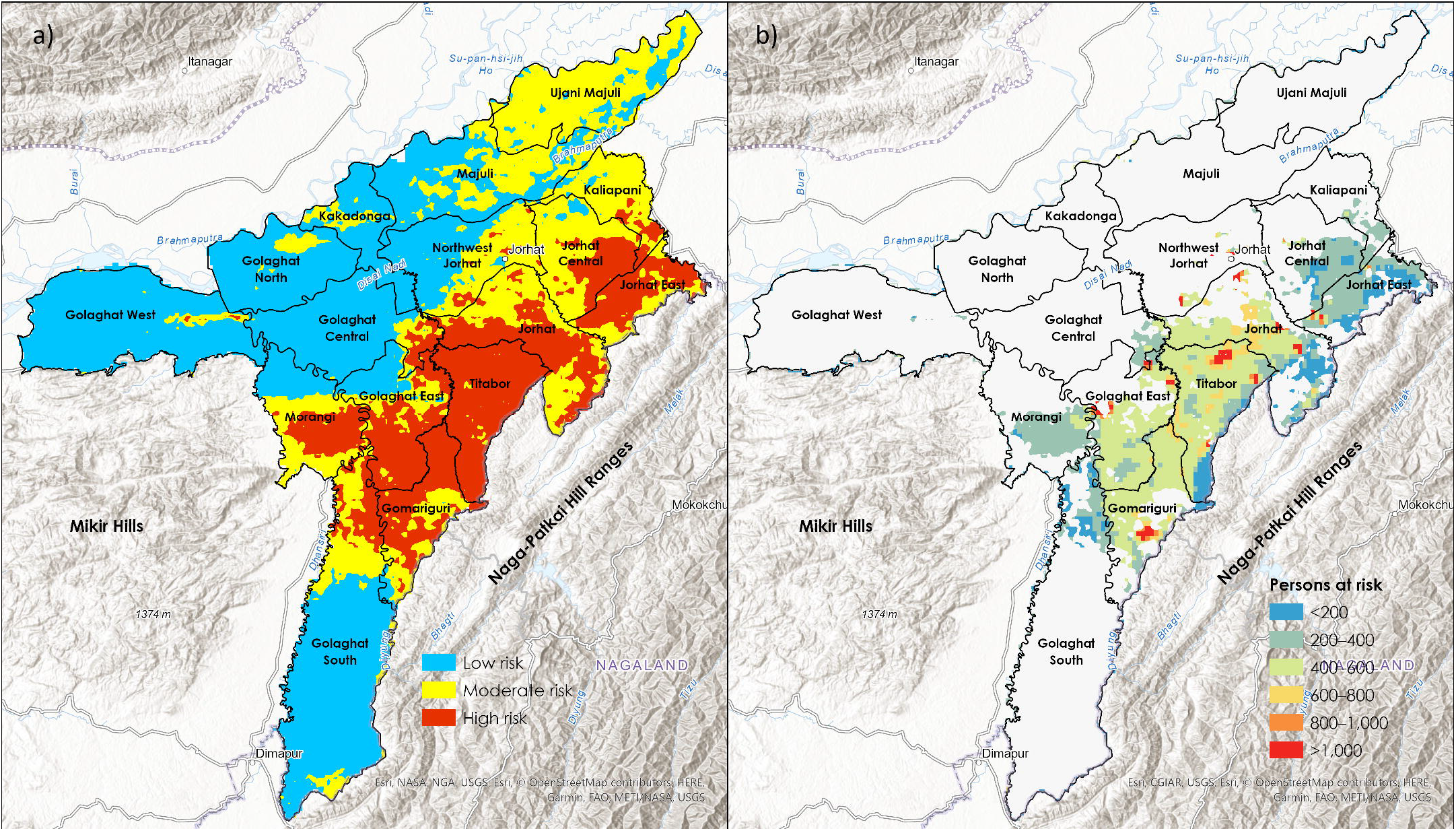
The total population (living in high and moderate risk areas), potentially exposed to As concentrations >10 µg/L in sixteen sub-districts of the study area.

## 5. Public health implications

The result of this study enables effective demarcation of the high and low-risk areas in two worst-hit districts of Assam, including potentially moderate-risk areas in the district of Majuli, where the inhabitants are relatively poor. The hazard probability map and risk model of the occurrence of As concentrations in groundwater exceeding the WHO guidelines of 10 µg/L will be helpful for the policymakers to manage the aquifer sustainably since agricultural use of groundwater is negligible in comparison with As-contaminated areas elsewhere in Bangladesh. In the short term, local administration can target the regions with the greatest risk (probability >0.70) and advice the local inhabitants regarding the potential danger of consuming high As drinking water. A targeted sampling campaign could help identify wells and regions and inform inhabitants whether they need to switch their drinking water sources to a safer tube well. In the long term, policymakers should target the exposed region by implementing additional water treatment plants with support from the Government of India or other sources.

Based on the analysis of piped water supply schemes (PWSS), we identified that high-risk sub-districts Gomariguri, Morangi, Golaghat East, and Golaghat South in Golaghat district are least connected to household tap waters (Fig. 8). Likewise, Titabor, Jorhat East, and Jorhat sub-districts in Jorhat districts are least connected to household tap waters provided by the government. This suggests a greater risk of As exposure due to the lack of safe water access in rural households in the high-risk areas. It would be important to generate awareness among the public to get access to this safe drinking water household connection. The local government could take the initiative by subsidizing the costs associated with getting these connections since many rural inhabitants might not be able to afford the costs. This will enable the local governments to accurately address the public health concerns and control the use of groundwater by discouraging the installation of private tube wells.

**Figure 8.**
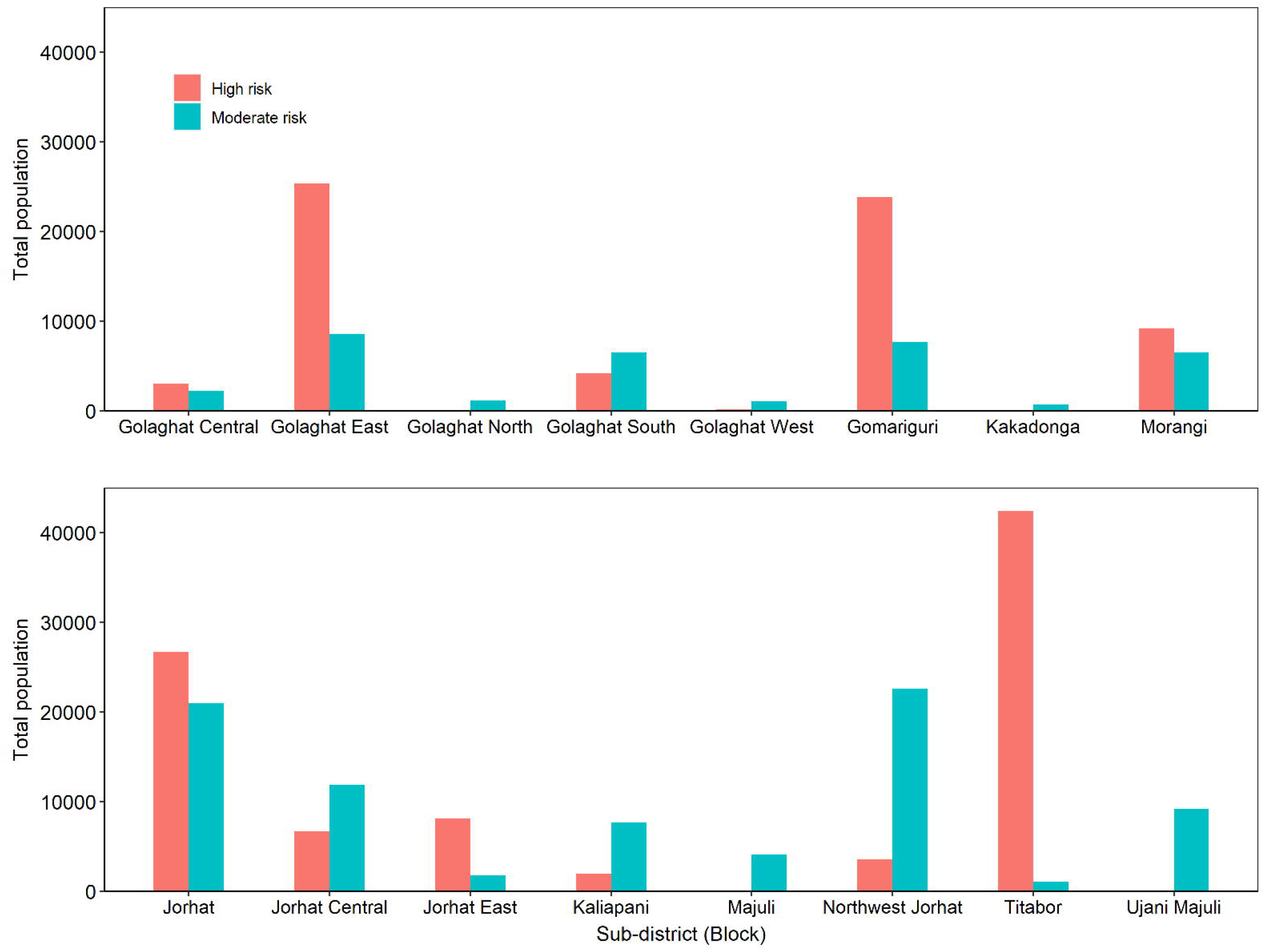
Percentage of the total population has either safe water access or no safe water access in their households in sixteen sub-districts of the study area.

## 6. Conclusion

The hazard model presented in this study provides an insight into the location and area where the inhabitants were potentially exposed to elevated As concentrations in groundwater. The hazard maps could be used as a guide in identifying targeted testing locations and assessing public health impacts. The hazard map is also relevant to finding appropriate locations to install community drinking water wells and treatment facilities to provide safe water access in potentially high-risk regions. The hazard map can also be used to inform villagers and generate community awareness about the potential impact of elevated As in groundwater through active participation.

The availability of groundwater As testing data from newer areas would undoubtedly help improve the model, particularly from Morangi, Golaghat South, Jorhat Central, and Majuli, where the data is almost absent for training the model. The hazard model presented here does not account for the role of aquifer depths in relation to the spatial pattern of As in the aquifer. In general, As concentration was found to vary with sediment age and depth of the tube wells. In addition, time could be incorporated into the model since As concentrations in tube wells changed with seasons. This is especially true for the tube wells located close to rivers with large fluctuations in post- and pre-monsoonal groundwater levels. A spatio-temporal model could provide significant insights into understanding aquifer contamination.

## Data Availability

All data produced in the present work are contained in the manuscript. The details are provided in Table 1.

## Notes

### Competing Interest Statement

The authors have declared no competing interest.

### Funding Statement

This study is funded by NASA grant number 80NSSC20K1718.

## References

1. Ahmed, K. M., Bhattacharya, P., Hasan, M. A., Akhter, S. H., Alam, S. M. M., & Bhuyian, M. A. H. (2004). Arsenic contamination in groundwater of alluvial aquifers in Bangladesh: An overview. Applied Geochemistry, 19(2), 181–200.

2. Amini, M., Abbaspour, K.C., Berg, M., Winkel, L., Hug, S.J., Hoehn, E., Yang, H., Johnson, C.A. Statistical modeling of global geogenic arsenic contamination in groundwater. Environ. Sci. Technol. 2008. 42, 3669–3675.

3. Ayotte, J.D., Nolan, B.T., Nuckols, J.R., Cantor, K.P., Robinson, G.R., Baris, D., Hayes, L., Karagas, M., Bress, W., Silverman, D.T., Lubin, J.H., 2006. Modeling the probability of arsenic in groundwater in New England as a tool for exposure assessment. Environ. Sci. Technol. 40, 3578–3585. https://doi.org/10.1021/es051972f.

4. Bangladesh Bureau of Statistics (BBS) and United Nations Children’s Fund (UNICEF). Bangladesh Multiple Indicator Cluster Survey 2012–2013. Progotir Pathey. Key District Level Findings. Dhaka, Bangladesh: BBS and UNICEF Bangladesh; 2014.

5. Bindal, S.; Singh, C.K. Predicting groundwater arsenic contamination: Regions at risk in highest populated state of India. Water Res. 2019, 159, 65–76.

6. Borah, K., Bhuyan, B., & Sarma, H. P. (2009). Lead, arsenic, fluoride, and iron contamination of drinking water in the tea garden belt of Darrang district, Assam, India. Environmental Monitoring and Assessment, 169(1-4), 347–352.

7. Breiman, L. Random forests. Mach. Learn. 2001, 45, 5–32.

8. Chakraborti, D., Rahman, M.M., Paul, K., Chowdhury, U.K., Sengupta, M.K., Lodh, D., Chanda, C.R., Saha, K.C., Mukherjee, S.C., 2002. Arsenic calamity in the Indian subcontinent what lessons have been learned? Talanta 58 (1), 3–22.

9. Choudhury, R., Nath, B., Khan, M.R., Mahanta, C., Ellis, T., van Geen, A., 2018. Impact of aquifer flushing across a 35 Km transect perpendicular to the upper Brahmaputra river in Assam, India. Water Resour. Res. 10.1029/2017WR022485

10. CGWB (2008). Central Ground Water Board, Ministry of Water Resources Government of India, Information Booklet. Jorhat District. Retrieved from http://cgwb.gov.in/

11. Chetia, M., Chatterjee, S., Banerjee, S., Nath, M. J., Singh, L., Srivastava, R. B., & Sarma, H. P. (2011). Groundwater arsenic contamination in the Brahmaputra river basin: A water quality assessment in the Golaghat (Assam), India. Environmental Monitoring and Assessment, 173(1-4), 371–385.

12. DPHE, 2001. Arsenic contamination of groundwater in Bangladesh. Dept. of Public Health Engineering, British Geological Survey, and Mott MacDonald. BGS Technical Report WC/00/19 (4 volumes).

13. Enmark, G., Nordborg, D., 2007. Arsenic in the groundwater of the Brahmaputra floodplains, Assam, India – Source, distribution and release mechanisms. Minor Field Study 131, Committee of Tropical Ecology, Uppsala University, Uppsala, Sweden, ISSN 1653-5634, 35p.

14. Farr, T. G., and M. Kobrick, 2000, Shuttle Radar Topography Mission produces a wealth of data. Eos Trans. AGU, 81:583–583.

15. Fawcett, T. An introduction to ROC analysis. Pattern Recognit. Lett. 2006, 27, 861–874.

16. Fick, S.E. and R.J. Hijmans, 2017. WorldClim 2: new 1km spatial resolution climate surfaces for global land areas. International Journal of Climatology 37 (12): 4302–4315.

17. Flanagan SV, Johnston RB, Zheng Y. Arsenic in tube well water in Bangladesh: health and economic impacts and implications for arsenic mitigation. Bull World Health Organ. 2012;90(11):839–846.

18. Foster, S.S.D.; Chilton, P.J. Groundwater: the processes and global significance of aquifer degradation. Phil. Trans. R. Soc. Lond. B 2003, 358, 1957–1972.

19. Ho, T.K. Random decision forests. In Proceedings of the 3rd International Conference on Document Analysis and Recognition, Montreal, QC, Canada, 14–16 August 1995; Volume 1. pp. 278–282.

20. Integrated Management Information System, 2021. Jal Jeevan Mission Reports. www.ejalshakti.gov.in/IMISReports/IMISReportLogin.aspx (accessed on 10 November 2021).

21. Hengl, T. Global DEM derivatives at 250 m, 1 km and 2 km based on the MERIT DEM (Version 1.0). Topographic Wetness Index, Zenodo, 5 October 2018.

22. Lehner, B., Verdin, K., Jarvis, A. (2008): New global hydrography derived from spaceborne elevation data. Eos, Transactions, AGU, 89(10): 93–94.

23. Mahanta, C., Choudhury, R., Basu, S., Hemani, R., Dutta, A., Barua, P. P., & Saikia, L. (2015). Preliminary assessment of arsenic distribution in Brahmaputra River basin of India based on examination of 56,180 public groundwater wells. In Safe and sustainable use of arsenic-contaminated aquifers in the Gangetic Plain (pp. 57–64). Guwahati, Assam: Springer Publishers.

24. McArthur, J. M., Banerjee, D. M., Hudson-Edwards, K. A., Mishra, R., Purohit, R., Ravenscroft, P., et al. (2004). Natural organic matter in sedimentary basins and its relation arsenic in anoxic ground water: The example of West Bengal and its worldwide implications. Applied Geochemistry, 19(8), 1255–1293.

25. McArthur, J. M., Nath, B., Banerjee, D. M., Purohit, R., & Grassineau, N. (2011). Palaeosol control on groundwater flow and pollutant distribution: The example of arsenic. Environmental Science and Technology, 45(4), 1376–1383.

26. Mukherjee, A., Sarkar, S., Chakraborty, M., Duttagupta, S., Bhattacharya, A., Saha, D., Bhattacharya, P., Mitra, A., Gupta, S., 2021. Occurrence, predictors and hazards of elevated groundwater arsenic across India through field observations and regional-scale AI-based modeling. Sci. Tot. Environ. 759, 143511.

27. Nath, B., Berner, Z., Basu Mallik, S., Chatterjee, D., Charlet, L., & Stueben, D. (2005). Characterization of aquifers conducting groundwaters with low and high arsenic concentrations: A comparative case study from West Bengal, India. Mineralogical Magazine, 69(05), 841–854.

28. Nath, B., Chakraborty, S., Burnol, A., Stueben, D., Chatterjee, D., Charlet, L., 2009. Mobility of arsenic in the sub-surface environment: An integrated hydrogeochemical study and sorption model of the sandy aquifer materials. J. Hydrol. 364, 236–248.

29. Pedregosa, F., et al. (2011) Scikit-Learn: Machine Learning in Python. Journal of Machine Learning Research, 12, 2825–2830.

30. Podgorski, J.E.; Labhasetwar, P.; Saha, D.; Berg, M. Prediction modeling and mapping of groundwater fluoride contamination throughout India. Environ. Sci. Technol. 2018, 52, 9889–9898.

31. Podgorski, J.; Wu, R.; Chakravorty, B.; Polya, D.A. Groundwater Arsenic Distribution in India by Machine Learning Geospatial Modeling. Int. J. Environ. i>Res. Public Health 2020a, 17, 7119. https://doi.org/10.3390/ijerph17197119

32. Podgorski, J., Berg, M., 2020b. Global threat of arsenic in groundwater. Science 368, 845–850.

33. Poggio, L., de Sousa, L. M., Batjes, N. H., Heuvelink, G. B. M., Kempen, B., Ribeiro, E., and Rossiter, D., 2021. SoilGrids 2.0: producing soil information for the globe with quantified spatial uncertainty. Soil, 7, 217–240.

34. Ravenscroft, P., Brammer, H., Richards, K.S., 2009. Arsenic pollution: a global synthesis. Wiley-Blackwell, Oxford, UK.

35. Rahman, M.M., Asaduzzaman, M., Naidu, R., (2011). Arsenic exposure from rice and water sources in the Noakhali district of Bangladesh. Environ. Geochem. Health 3, 1–10.

36. Rodríguez, R., Ramos, J.A., Armienta, A., 2004. Groundwater arsenic variations: the role of local geology and rainfall. Appl. Geochem. 19, 245–250.

37. Roy, P.S., P. Meiyappan, P.K. Joshi, M.P. Kale, V.K. Srivastav, S.K. Srivasatava, M.D. Behera, A. Roy, Y. Sharma, R.M. Ramachandran, P. Bhavani, A.K. Jain, and Y.V.N. Krishnamurthy. 2016. Decadal Land Use and Land Cover Classifications across India, 1985, 1995, 2005. ORNL DAAC, Oak Ridge, Tennessee, USA. https://doi.org/10.3334/ORNLDAAC/1336

38. Ryo, M., Rillig, M.C., 2017. Statistically reinforced machine learning for nonlinear patterns and variable interactions. Ecosphere https://doi.org/10.1002/ecs2.1976

39. Saha, K.C., 1995. Chronic arsenic dermatosis from tubewell water in West Bengal during 1983– 87. Ind. J. Dermatol. 40, 1–12.

40. Shamsudduha, M., Uddin, A., Saunders, J.A., Lee, M.K., 2008. Quaternary stratigraphy, sediment characteristics and geochemistry of arsenic-contaminated alluvial aquifers in the ganges-brahmaputra floodplain in Central Bangladesh. J. Contam. Hydrol. 99 (1-4), 112–136.

41. Singh, A.K., 2004. Arsenic contamination in groundwater of North Eastern India. Proc. 11th National Symposium on Hydrology with focal theme on Water Quality, National Institute of Hydrology, Roorkee, India, 255–262.

42. Smith AH, Lingas EO, Rahman M (2000) Contamination of drinking-water by arsenic in Bangladesh: a public health emergency. Bull World Health Organ 78:1093–1103

43. Sturges, H.A. The choice of a class interval. J. Am. Stat. Assoc. 1926, 21, 65–66.

44. Trabucco, A.; Zomer, R.J. Global Aridity Index (Global-Aridity) and Global Potential Evapo-Transpiration (Global-PET) Geospatial Database; CGIAR Consortium for Spatial Information. 2009. CGIAR-CSI GeoPortal. Available online: http://www.cgiar-csi.org (accessed on 10 November 2021).

45. van Geen, A., Zheng, Y., Versteeg, R., Stute, M., Horneman, A., Dhar, R.K., Steckler, R., Gelman, M., Small, C., Ahsan, H., Graziano, J.H., Hussein, I., Ahmed, K.M., 2003. Spatial variability of arsenic in 6000 tubewells in a 25 km^2^ area of Bangladesh. Water Resour. Res. 39, 1140.

46. Verma, S., Mukherjee, A., Mahanta, C., Choudhury, R., & Mitra, K. (2016). Influence of geology on groundwater-sediment interactions in varied arsenic enriched tectono-morphic aquifers of the Brahmaputra River basin. Journal of Hydrology, 540, 176–195.

47. Wu, R., Podgorski, J., Berg, M., Polya, D.A., 2021. Geostatistical model of the spatial distribution of arsenic in groundwaters in Gujarat State, India. Environ. Geochem. Health 43, 2649–2664.

48. www.worldpop.org

